# EVALUATION OF ELEVEN IMMUNOCHROMATOGRAPHIC ASSAYS FOR SARS-CoV-2 DETECTION: INVESTIGATING DENGUE CROSS-REACTION

**DOI:** 10.1101/2020.10.09.20210039

**Authors:** Beatriz Araujo Oliveira, Lea Campos de Oliveira, Franciane Mendes de Oliveira, Geovana Maria Pereira, Regina Maia de Souza, Erika Regina Manuli, Fabricio Klerynton Marchini, Evelyn Patrícia Sanchez Espinoza, Marcelo Park, Leandro Taniguchi, Pedro Vitale Mendes, Lucas Augusto Moyses Franco, Ana Catharina Nastri, Maura Salaroli de Oliveira, José Mauro Vieira Junior, Esper Georges Kallas, Anna Sara Levin, Ester Cerdeira Sabino, Silvia Figueiredo Costa

## Abstract

**Background:** COVID-19 disease (Coronavirus disease 2019) caused by SARS-CoV-2 (Severe acute respiratory syndrome coronavirus 2) is widespread worldwide, affecting more than 11 million people globally (July 6^th^, 2020). Diagnostic techniques have been studied in order to contain the pandemic. Immunochromatographic (IC) assays are feasible and low cost alternative for monitoring the spread of COVID-19 in the population.

**Methods:** Here we evaluate the sensitivity and specificity of eleven different immunochromatographic tests in 98 serum samples from confirmed cases of COVID-19 through RT-PCR and 100 negative serum samples from blood donors collected in February 2019. Considering the endemic situation of Dengue in Brazil, we also evaluated the cross-reactivity with Dengue using 20 serum samples from patients with confirmed diagnosis for Dengue collected in early 2019 through four different tests.

**Results:** Our results demonstrated agreement between immunochromatographic assays and RT-PCR, especially after 10 days since the onset of symptoms. The evaluation of IgG and IgM antibodies combined demonstrated a strong level of agreement (0.85) of IC assays and RT-PCR. It was observed cross-reactivity between Dengue and COVID-19 using four different IC assays for COVID-19 diagnosis. The specificity of IC assays to detected COVID-19 IgM antibodies using Dengue serum samples varied from 80% to 85%; the specificity of IgG detection was 100% and total antibody was 95%.

**Conclusions:** We found high sensitivity, specificity and good agreement of IC assays, especially after 10 days onset of symptoms. However, we detected cross-reactivity between Dengue and COVID-19 mainly with IgM antibodies demonstrating the need for better studies about diagnostic techniques for these diseases.

**Highlights:**

- Immunochromatographic assays demonstrated high sensitivity and specificity and good agreement with the gold-standard RT-PCR;
- Increase in sensitivity and specificity of assays using samples collected after the 10^th^ day of symptoms;
- Cross-reaction with Dengue serology in evaluation of IgM.

## 1. Introduction

COVID-19 (Coronavirus disease 2019) is a human infectious disease caused by a new betacoronavirus SARS-Cov-2 or 2019-nCoV (Severe acute respiratory syndrome coronavirus 2) firstly reported in China by flu-like symptoms (December 26^th^, 2019) and now widespread worldwide, affecting more than 11 million people globally according to World Health Organization (July 6^th^, 2020) [1–5].

The COVID-19 diagnosis is based on clinical and epidemiological features, image exams and finally the analysis of nucleic acids through reverse-transcription polymerase chain reaction (RT-PCR), established as the gold standard for COVID-19 disease [1,2,6]. However, the accuracy of this method depends on the viral load on the collection site and on time from symptom onset. It presents limitations such as incorrect collection and processing of samples, the need of expensive equipment and reagents, trained operators and the delay in releasing the results [1,2,6,7].

The immunochromatographic (IC) assay is a feasible and low-cost alternative for monitoring the spread of COVID-19 in the population in general and specially in low and middle-income countries which lack of structure. Contact-tracing, population survey including health workers, teachers and students on resuming classes are situations that may be useful to use serology since the sensitivity of RT-PCR in asymptomatic is low and ranges from 8 to 10% [1,2].

Likewise COVID-19, Dengue is also an emerging disease, especially in tropical and subtropical countries transmitted by Aedes mosquito, and both diseases are similar regarding clinical and laboratory features hampering the diagnosis [8–10]. Another matter is the temporal dynamic of both diseases in Brazil. The spread of Dengue usually increases between March and April due the rainy seasons, the same period when respiratory diseases are most common, and period in which the number of COVID-19 confirmed cases started to rise [10].

Despite the high number of IC assays available, the readiness to perform the diagnosis and advantages involving its use, here we evaluate the sensitivity and specificity of eleven different IC assays in serum samples from confirmed cases of COVID-19 through RT-PCR and negative serum samples from blood donors collected in February 2019. Considering the endemic situation of Dengue in Brazil, we also evaluated the cross-reactivity with Dengue using serum samples from patients with confirmed diagnosis for Dengue collected in early 2019 through four different tests.

## 2. Methods

### 2.1 Study design

This is a prospective multicenter study of COVID-19 hospitalized patients at two Brazilian Hospitals: Hospital das Clínicas da Faculdade de Medicina da Universidade de São Paulo (HC-FMUSP), a public teaching hospital with 2,000 beds; and Hospital Sírio-Libanes (HSL), a private 400-bed hospital. Both hospitals are located in Sao Paulo.

### 2.2 Ethical approval

This study was approved by the Brazilian national ethics review board (CONEP), protocol number 30701920200000068.

### 2.3 Samples collection

The positive serum samples used in this study were collected in April at HSL and HC-FMUSP from 74 symptomatic patients with confirmed COVID-19 diagnosis through RT-PCR, besides these 74 samples other 24 samples were collected from patients with more than 10 days of symptoms, altogether 98 positive samples were used in this study. The serum samples were stored at −20 until the moment of test. In addition, 100 serum samples collected in February 2019 from blood donors at Fundação Pró-sangue – Hemocentro de São Paulo (São Paulo, Brazil) were used as negative controls for experiments, these samples were also stored at −20 until the moment of test. To evaluate the cross reactivity with Dengue we analyzed 20 samples collected from patients with confirmed diagnosis for Dengue by ELISA technique between February and March/2019.

### 2.4 Immunochromatographic assays for antibodies against SARS-CoV-2

In this study positive and negative samples from Brazilian subjects were evaluated through eleven qualitative IC assays performed according to manufacturer instructions summarized at table 1, using the following kits: One Step COVID-19 test (Wondfo, China – batch W19500341); SARS-COV-2 (COVID-19) IgG/IgM Antibody Fast Detection (Thermogenesis, China – batch SYG202010); SARS-COV-2 IgM/IgG Antibody Test Kit (Luxus, China – batch SYG202010); COVID-19 IgM/IgG Rapid Test Kit (Camtech, Singapore – batch NF3170); Rapid test COVID-19 IgG/IgM BIO (Bioclin, Brazil – batch 0010); SARS-CoV-2 IgG / IgM Rapid Test Kit (TBG, Taiwan – batch FRS20041K); COVID-19 IgG/IgM ECO TEST (Ecotest, Brazil – batch 202005043); MedTest Coronavírus (COVID) IgG/IgM (MedTest, China – batch COV20030081); SARS-Cov-2 antibody detection test (Lepu, China – batch 20CG2518X); Kit COVID-19 IgG/IgM LF (Advagen, Brazil – batch L20183-02) and COVID-19 (SARS-CoV-2) IgM/IgG antibody Test kit (MedNet Wuhan, China – batch 20030501).

**Table 1.** Instructions of eleven immunochromatographic (colloidal gold) tests

| Label | Test name | Manufacturer country | Detection | Amount of Sample | Amount of Reagent | Incubation period |
| --- | --- | --- | --- | --- | --- | --- |
| Advagen | Kit COVID-19 IgG/IgM LF | Brazil | IgG/IgM | 10 µl | 40 µl | 15 minutes |
| Bioclin | Rapid test COVID-19 IgG/IgM BIO | Brazil | IgG/IgM | 10 µl | 2 drops (60~80 µl) | 10 ~ 15 minutes |
| CamTech | COVID-19 IgM/IgG Rapid Test Kit | Singapore | IgG/IgM | 10 µl | 2 drops (60 µl) | 15 minutes |
| Ecotest | COVID-19 IgG/IgM ECO TEST | Brazil | IgG/IgM | 10 µl | 3 drops (90 µl) | 10 ~ 15 minutes |
| Lepu | SARS-Cov-2 antibody detection test | China | IgG/IgM | 10 µl | 2 drops (80 µl) | 10 ~ 20 minutes |
| Luxus | SARS-COV-2 IgM/IgG Antibody Test Kit | China | IgG/IgM | 20 µl | 80 µl | 3 minutes |
| MedNet Wuhan | COVID-19 (SARS-CoV-2) IgM/IgG antibody Test kit | China | IgG/IgM | 10 µl | 2 drops (70 µl) | 15 minutes |
| MedTest | MedTest Coronavirus (COVID) IgG/IgM | China | IgG/IgM | 10 µl | 2 drops (80 µl) | 10 minutes |
| TBG | SARS-CoV-2 IgG / IgM Rapid Test Kit | Taiwan | IgG/IgM | 10 µl | 2 drops | 15 minutes |
| Thermogenesis | SARS-COV-2 (COVID-19) IgG/IgM Antibody Fast Detection | China | IgG/IgM | 20 µl | 80 µl | 3 minutes |
| Wondfo | One Step COVID-19 test | China | Total IG | 10 µl | 2~3 drops (80 µl) | 15 ~ 20 minutes |

### 2.5 Cross-reactivity between Dengue and COVID-19 serology

To evaluate the cross-reactivity of COVID-19 disease and Dengue serology, 20 samples of confirmed Dengue cases diagnosed through ELISA technique on early 2019 were evaluated using four IC assays for detection of antibodies against SARS-CoV-2: One Step COVID-19 test (Wondfo, China – batch W19500341), COVID-19 IgM/IgG Rapid Test Kit (Camtech, Singapore – batch NF3170), Kit COVID-19 IgG/IgM LF (Advagen, Brazil – batch L20183-02) and IgM/IgG antibody Test kit (MedNet Wuhan, China – batch 20030501) according to manufacturer instructions. In addition, 40 samples from COVID-19 confirmed cases were evaluated using PANBIO DENGUE IgM CAPTURE ELISA (Abbott, USA – batch 01P20E014) in which 31 of them were collected after 10 days of symptoms. We also evaluated the cross-reactivity using IC assays for detection of antibodies against Dengue in 33 samples of COVID-19 confirmed cases using ALERE DENGUE DUO-NS1 IgG and IgM (Abbott, USA – batch 11DDE008A-A and 11DDE007A-A).

### 2.6 Statistical analysis

The validity of tests was measured through sensitivity (true positive / true positive + false negative = %) and specificity (true negative / true negative + false positive = %) calculations [11]. The concordance analysis was performed by the Fleiss’s Kappa method comparing the agreement between the gold standard (RT-PCR) and each one of the eleven-tested IC assays, the confidence interval was 95%.

## 3. Results

Altogether 198 serum samples were collected for this study, among them, 98 samples were collected from confirmed COVID-19 patients through RT-PCR, 74 of these patients’ samples were collected before the 10^th^ day since the onset of symptoms and 24 samples were collected after 10 days since the beginning of symptoms. In addition, as negative control were used 100 samples collected from blood donors on early 2019. The sensitivity and specificity as well as agreement ratio of eleven commercial qualitative IC assays were evaluated; the results were obtained through tests performed between May/2020 and July/2020.

### 3.1 Sensitivity of IC assay

A panel containing 74 positive samples and 100 negative samples was used to evaluate the sensibility of eleven IC tests. As summarized in table 2, among the tests, ten of them describe separately the detection of IgG and IgM antibodies and one of them Wondfo (China) evidence the presence of total immunoglobulins.

**Table 2.** Quality measurements of immunochromatographic assays

| Label | Detection | Nº of positive samples | Nº of negative samples | Sensitivity | Specificity | Fleiss's Kappa (95% CI) |
| --- | --- | --- | --- | --- | --- | --- |
| Advagen | IgG | 50 | 50 | 75.2% | 100% | 0.74 (0.54~0.93) |
| Advagen | IgM | 50 | 50 | 35.5% | 96.7% | 0.23 (0.03~0.42) |
| Bioclin | IgG | 49 | 50 | 81% | 100% | 0.82 (0.62~1.01) |
| Bioclin | IgM | 49 | 50 | 61% | 98% | 0.58 (0.38~0.78) |
| CamTech | IgG | 40 | 40 | 70% | 100% | 0.69 (0.47~0.91) |
| CamTech | IgM | 40 | 40 | 87.5% | 97.5% | 0.85 (0.63~1.07) |
| Ecotest | IgG | 20 | 20 | 90% | 100% | 0.58 (0.27~0.89) |
| Ecotest | IgM | 20 | 20 | 90% | 100% | 0.58 (0.27~0.89) |
| Lepu | IgG | 10 | 10 | 90% | 100% | 0.90 (0.46~1.34) |
| Lepu | IgM | 10 | 10 | 90% | 100% | 0.90 (0.46~1.34) |
| Luxus | IgG | 40 | 40 | 75% | 100% | 0.75 (0.53~0.97) |
| Luxus | IgM | 40 | 40 | 72.5% | 100% | 0.72 (0.50~0.94) |
| MedNet Wuhan | IgG | 40 | 40 | 77.5% | 97.5% | 0.72 (0.50~0.94) |
| MedNet Wuhan | IgM | 40 | 40 | 80% | 95% | 0.75 (0.53~0.97) |
| MedTest | IgG | 15 | 10 | 80% | 100% | 0.76 (0.37~1.15) |
| MedTest | IgM | 15 | 10 | 93.3% | 100% | 0.83 (0.43~1.23) |
| TBG | IgG | 15 | 10 | 93.3% | 100% | 0.84 (0.45~1.23) |
| TBG | IgM | 15 | 10 | 80% | 100% | 0.76 (0.37~1.15) |
| Thermogenesis | IgG | 40 | 40 | 80% | 100% | 0.80 (0.58~1.02) |
| Thermogenesis | IgM | 40 | 40 | 80% | 100% | 0.80 (0.58~1.02) |
| Wondfo | Total IG | 74 | 100 | 76.7% | 100% | 0.78 (0.63~0.93) |

The greater sensitivity for detection of only IgG antibody among ten IC tests was observed in TBG (Taiwan) IC test at 93.3%, followed by Ecotest (Brazil) and Lepu (China) IC test at 90%, Bioclin (Brazil) at 81%, MedTest (China) and Thermogenesis (China) presented equivalent results (80%), followed by MedNet (China), Advagen (Brazil) and Luxus (China) at 77.5%, 75.2% and 75% respectively. The Camtech (Singapore) presented the lower sensitivity among the tests at 70%.

The sensitivity for detection of only IgM antibody was also evaluated, the better result was achieve by MedTest (China) at 93.3%, followed by Ecotest (Brazil) and Lepu (China) at 90%. The Camtech (Singapore) presented 87.5% of sensitivity while TBG (Taiwan), Thermogenesis (China) and MedNet (China) achieved identical sensitivities of 80%. Luxus (China) sensibility was 72.5% followed by Bioclin (Brazil) at 61% and finally Advagen (Brazil) at 35.5%. The evaluation of total immunoglobulins presented a sensitivity of 76.7% as observed in Wondfo test (China).

### 3.2 Specificity of IC assay

The assay specificity was evaluated according to table 2. In IgG IC assays the specificity was equal (100%) in all the tests except for MedNet (China) at 97.5%. In the evaluation of IgM detection the assays sensitivity was 100% for Ecotest (Brazil), Lepu (China), Luxus (China), MedTest (China), TBG (Taiwan) and Thermogenesis (China). Bioclin (Brazil) achieve a specificity of 98%, followed by Camtech (Singapore) at 97.5%, Advagen (Brazil) at 96.7% and MedNet (China) at 95%. The evaluation of total immunoglobulins presented a specificity of 100% for Wondfo (China).

### 3.3 Agreement rate

To evaluate the agreement among the testes and the gold standard RT-PCR we analyzed the Kappa’s value described in table 2, among the evaluated tests for detection of IgG antibody, four of them presented a strong agreement level: Lepu (China) 0.90, TBG (Taiwan) 0.84, Bioclin (Brazil) 0.82 and Thermogenesis (China) 0.80. Five tests presented a moderate agreement level MedTest (China) 0.76, Luxus (China) 0.75, Advagen (Brazil) 0.74, MedNet (China) 0.72 and Camtech (Singapore) 0.69. Finally, one presented a weak agreement level Ecotest (Brazil) 0.58.

The agreement of IgM detection tests was strong for four of evaluated tests: Lepu (China) 0.90, Camtech (Singapore) 0.85, MedTest (China) 0.83 and Thermogenesis (China) 0.80. A moderate agreement was observed in three tests: TBG (Taiwan) 0.76, MedNet (China) 0.75 and Luxus (China) 0.72. A weak agreement was observed in two tests: Ecotest (Brazil) and Bioclin (Brazil) 0.58, followed by a minimal agreement Advagen (Brazil) 0.23. The evaluation of total immunoglobulins demonstrated a moderate agreement level of Wondfo (China) 0.78.

### 3.4 Evaluation according to stage of disease

Considering the temporal dynamic of antibodies, we evaluated the sensitivity of tests in samples from patients collected more than 10 days after the onset of symptoms according to table 3. The sensitivity observed for IgG assays was 91.7% in MedNet (China) and 79.2% in Bioclin (Brazil) and a strong agreement level between these two tests and RT-PCR was note through Kappa value of 0.83. Evaluating the agreement ratio among MedNet (China) and RT PCR and Bioclin (Brazil) compared to RT-PCR the Kappa value of 0.87 and 0.79 was obtained respectively.

**Table 3.** Evaluation of samples collected more than 10 days after the beginning of symptoms

| Label | Detection | Sensitivity | Fleiss's Kappa |
| --- | --- | --- | --- |
| Bioclin | IgG | 79.2% | 0.79 (0.51~1.07) |
| MedNet Wuhan | IgG | 91.7% | 0.87 (0.59~1.16) |
| Fleiss's Kappa: 0.83 (0.67~1.0) |  |  |  |
| Bioclin | IgM | 70.1% | 0.66 (0.38~0.94) |
| MedNet Wuhan | IgM | 91.7% | 0.83 (0.55~1.12) |
| Fleiss's Kappa: 0.75 (0.58~0.91) |  |  |  |
| Bioclin | IgG and IgM | 83.3% | 0.79 (0.51~1.07) |
| MedNet Wuhan | IgG and IgM | 91.7% | 0.79 (0.51~1.07) |
| Wondfo | Total IG | 95.8% | 0.92 (0.63~1.20) |
| Fleiss's Kappa: 0.85 (0.73~0.96) |  |  |  |

In IgM assays the MedNet (China) sensitivity was 91.7% while Bioclin (Brazil) sensitivity was 70.1%, the agreement ratio among these tests and RT-PCR was moderate Kappa value of 0.75, evaluating the agreement ratio between MedNet (China) and RT-PCR was obtained Kappa value of 0.83 and Bioclin (Brazil) presented a Kappa value of 0.66.

The evaluation of IgG and IgM antibodies combined demonstrated a strong level of agreement (0.85) of tests and RT-PCR results besides the sensitivity of 95.8%, 91.7% and 83.3% for Wondfo (China), MedNet (China) and Bioclin (Brazil) respectively. An evaluation of agreement between IgG and IgM tests and RT-PCR demonstrated an almost perfect agreement ratio for Wondfo (China), kappa value of 0.92, followed by moderate agreement ratio of Bioclin (Brazil) and MedNet (China), kappa value 0.79.

### 3.5 Cross-reactivity between Dengue and COVID-19 detection

Forty samples of COVID-19 confirmed cases were evaluated using tests for detection of antibodies against Dengue, the specificity of the ELISA PANBIO DENGUE CAPTURE ELISA (Abbott, USA) achieve 100% specificity. The IC assay ALERE DENGUE DUO-NS1 IgG and IgM (Abbott, USA) demonstrated 100% specificity for both IgM and IgG antibodies using 33 samples of COVID-19 patients.

In the analyze of immunochrographic tests for COVID-19 diagnosis Advagen (Brazil) and Camtech (Singapore) reached 100% specificity for IgG antibody detection using 20 dengue samples while MedNet (China) achieved 95%. In IgM detection the specificity was 85% for Advagen (Brazil) and Camtech (Singapore), MedNet (China) reached 80% specificity while Wondfo specificity was 95% for detection of total antibodies as described in table 4.

**Table 4.** Evaluation of cross-reactivity between Dengue and COVID-19

| Label | Detection | Number of samples | Specificity |
| --- | --- | --- | --- |
| PANBIO DENGUE ELISA | IgM | 40 | 100% |
| NSF1 Dengue | IgG | 33 | 100% |
| NSF1 Dengue | IgM | 33 | 100% |
| Advagen | IgG | 20 | 100% |
| Advagen | IgM | 20 | 85% |
| Camtech | IgG | 20 | 100% |
| Camtech | IgM | 20 | 85% |
| MedNet | IgG | 20 | 95% |
| MedNet | IgM | 20 | 80% |
| Wondfo | Total IG | 20 | 95% |

## 4. Discussion

In this study, we evaluated eleven IC colloidal-gold qualitative based assays and analyzed the quality measurements of each one of them. According to our results, most of IC assays provide high sensitivity and specificity compared to the gold standard RT-PCR, especially after 10 days since the onset of disease with an excellent agreement ratio compared to the gold standard using Wondfo test evaluating total antibodies against SARS-CoV-2.

The COVID-19 pandemic impacted the world health system in an unprecedented way. In Brazil the first reported case occurred on February 27^th^, 2020, since them more than 1 million Brazilian people tested positive for COVID-19 [4,12]. Besides COVID-19, Brazil also faces an endemic situation regarding to Dengue, and considering the importance of both diseases, here we evaluated the cross-reactivity of Dengue and COVID-19 using different tests.

According to our results, the specificity in detection of IgG antibodies was greater in comparison with IgM detection using IC assays for detection of COVID-19 in Dengue patient’s samples. It is widely known that before the high-affinity response by IgG antibodies to pathogens the first defense provided by organism occurs through IgM molecules [13]. Our results suggest that the evaluation of total antibodies (IgG and IgM) is a useful tool to broaden the range of detection, enabling the evaluation of acute phase through IgM detection and convalescent phase by the presence of IgG. The evaluation of combined antibodies helps not only the discrimination of Dengue and COVID-19 but also avoid false negative results for COVID-19 and contributes to the establishment of control measures. On the other hand, the use of IC assays that detect IgM may have cross-reaction with Dengue and lead to a false positive result that might impact on the clinical management of the patient.

The IC assay is based on the detection of antibodies secreted by B lymphocytes against the viral pathogens using serum, plasma or whole blood samples from patients and a specific buffer on an immunochromatographic cassette [1,2,14,15]. The cassette is composed by plastic backing involving the strip where reaction occurs, sample pad for sample addition, conjugated pad containing antigens bound to colloidal nanoparticles, absorbent pad for waste reservoir, and the nitrocellulose membrane in which by capillary attraction the antigen-antibody reaction occurs, evidenced through a colored band (red color for colloidal-gold nanoparticles and blue color for colloidal selenium nanoparticles) in about 15 to 20 minutes [1,2,14,15].

Immunochromatographic is an easy and affordable method for diagnosis enabling prompt results in large scale. According to our results, the agreement ratio for detection of only IgG antibodies before the 10° day of symptom using IC technique in comparison with RT-PCR was moderate in the majority of used tests (50%), followed by strong agreement (40%) and weak agreement (10%). In contrast, for only IgM detection the agreement ratio between IC and RT-PCR was strong in the majority of tests (40%) followed by a moderate (30%), weak (20%) and minimal (10%) agreement. The evaluation of total antibodies using Wondfo test demonstrated better results using samples collected after the 10° day of symptoms (almost perfect agreement) than before this period (moderate agreement) compared to RT-PCR, the increase in sensitivity was also observed using MedNet test in samples after 10 days since the onset of symptoms.

In the literature, it is possible to find the detection of immunoglobulin on the 5^th^ day since the onset of disease. However, this detection is greater from the 8^th^ day on [15]. Our results are in accordance with literature, demonstrating an increase in sensitivity levels especially using the combined assessment of IgG and IgM antibodies.

Our study has limitations such as sample size; however, to our knowledge it was the first study that analyzed several SARS-CoV-2 IC tests and cross-reaction with Dengue. Thus, our findings can be useful in countries with high prevalence of Dengue to alert to the possible cross-reactivity of the SARS-CoV-2 IC assays, mainly IgM, with Dengue antibodies.

## 5. Conclusion

We found in general high sensitivity and specificity and good agreement of IC assays, especially after 10 days since the onset of symptoms. Our results also evidenced the importance in evaluating total immunoglobulins to increase sensitivity and specificity of IC assays. On the other hand, we detected cross-reactivity between Dengue and COVID-19 demonstrating the need for better studies and improvements in diagnostic techniques for these diseases.

## 6. Limitations of the study

In this study, a small number of samples was tested due to the availability of tests that we had at the beginning of COVID-19 in Brazil. Studies with more samples are needed to ensure validity of the results.

## Data Availability

All data are available in the manuscript attached in pdf format.

## 7. Author contribution

Oliveira, BA; Oliveira LC: Conceptualization, methodology, formal analysis and writing – original draft. Both authors contributed equally in the development of the study.

Oliveira FM; Pereira, GM; Souza RM: Validation and investiagation.

Manuli, ER; Marchini, FK; Park, Marcelo; Taniguchi, L; Mendes, PV; Franco, LAM; Espinoza, EPS; Nastri, AC; Levin, AS; Oliveira, MS; Vieira Junior, JM; Kallas, EG: Resources.

Costa, SF; Sabino, EC: Conceptualization, supervision and writing – review & editing. Both authors contributed equally in the development of the study.

